# Systematic review and meta-analysis of the prevalence of coronavirus in the world: One health approach is urgent

**DOI:** 10.1101/2021.06.09.21258651

**Authors:** Ricardo Faustino, Miguel Faria, Mónica Teixeira, Filipe Palavra, Maria do Céu Costa, Paulo Sargento

## Abstract

Coronaviruses have been responsible for major epidemic crises in 2003 with SARS-CoV-1, in 2012 with MERS-CoV and in 2019 with SARS-CoV-2 (COVID-19), causing serious atypical pneumonia in humans. We intend, with this systematic analysis and meta-analysis, to clarify the prevalence of the various strains of coronavirus in different animal species. For this purpose, we carried out an electronic survey using Pubmed’s Veterinary Science search tool to conduct a systematic assessment of published studies reporting the prevalence of different strains of coronavirus in different animal species between 2015 and 2020. We conducted different analysis to assess sensitivity, publication bias, and heterogeneity, using random and fixed effects. The final meta-analysis included 42 studies for systematic review and 29 in the meta-analysis. For the geographic regions with a prevalence greater than or equal to 0.20 (Forest plot overall; prevalence = 0.20, p < 0.01, Q = 10476.22 and I2 = 100%), the most commonly detected viruses were: enteric coronavirus (ECoV), pigeon-dominant coronavirus, (PdCoV), Avian coronavirus M41, Avian coronavirus C46, Avian coronavirus A99, Avian coronavirus JMK, MERS-CoV, Bovine coronavirus, Ro-BatCoV GCCDC1, Alphacoronavirus, Betacoronavirus, Deltacoronavirus, Gamacoronavirus and human coronaviruses (HCoVs). The wide presence of different strains of coronavirus in different animal species on all continents demonstrates the great biodiversity and ubiquity of these viruses.

The most recent epidemiological crises caused by coronavirus demonstrates our unpreparedness to anticipate and mitigate emerging risks, as well as the need to implement new epidemiological surveillance programs for viruses. Combined with the need to create advanced training courses in One Health, this is paramount in order to ensure greater effectiveness in fighting the next pandemics.

## Introduction

Coronavirus disease 2019 (COVID-19) is the most recent viral pandemic event in recent years associated with the *Coronoviridae* family, with COVID-19 being its seventh member [1]. The *Coronaviridae* comprise two subfamilies, including *Coronavirinae*, whose members are commonly referred to as coronaviruses (CoVs).

The outbreak was thought to have originated in Wuhan, spread rapidly to neighbouring provinces and, within three months, a pandemic was declared. Cases have been reported in every region of the world, with a high number of infections and deaths. New origin hypotheses,however, have been advanced in more recent studies [2].

Research studies indicate that 72% of events arising from zoonotic diseases originate from wildlife. Many of these diseases pose serious risks to human health, as demonstrated by the 2014 Ebola virus in West Africa, MERS-CoV in the Middle East in 2012 [3], SARS-CoV detected in 2002 in China and H5N1 in 2004. The existence of markets for trade of live animals brings wildlife closer to humans and domestic animals. These places of commerce have a potential role as an interface for the transmission of pathogens. This interface can contribute to the emergence of diseases, and the spread of a range of diseases, including pathogenic avian influenza H5N1, Severe Acute Respiratory Syndrome (SARS) [4,5]. According to Leroy [2], since genetic recombination events within human CoVs are well documented, the known high prevalence of dog infections with canine coronaviruses in Europe might foster recombination with SARS-CoV-2 if an animal were infected with both viruses. Such an event, if it happens, could lead to the emergence of a new coronavirus with unpredictable phenotypic characteristics (transmissibility and virulence). Unfortunately, the likelihood of such a scenario is difficult to assess.

The coronavirus is well known in the world of veterinary medicine. A translation of this experience can be very beneficial for human health [6,7]. For this purpose, One Health appears as an important concept. It is a new approach that is based on the relationship between humans, animals and the environment, and recognizes that the health and well-being of human beings is strongly related to the health of animals and their environment [3].

Belonging to the *Coronaviridae* family, in the order Nidovirales, the SARS-CoV-2 genome consists of positive-sense, single-stranded, polyadenylated, nonsegmented RNA [8]. In order to understand the importance and evolution of the coronavirus, a broader point of view is needed to understand the behaviour of *Coronaviridae*. To date, from the seven coronaviruses reported in humans, four of them are ubiquitous with seasonal circulation and mostly causing relatively mild colds (HKU1, NL63, OC43 and 229E). The other three, of more recent zoonotic origin, are associated with severe acute respiratory syndromes, namely SARS-CoV, MERS-CoV and now SARS-CoV-2. Of these seven human coronaviruses, NL63 and 229E belong to the alpha-CoV genus, while the other five are included within the beta-CoV genus. Coronaviruses detected in dogs and cats also belong to these two viral genera [9,10]. Like SARS-CoV-2 and the other respiratory syndrome viruses, the canine respiratory coronavirus (CRCoV), responsible for a respiratory condition in dogs, belongs to the beta-CoV genus. Canine coronavirus (CCoV) and Feline coronavirus (FCoV), both responsible for digestive diseases, belong to *Alphacoronavirus*.

Therefore, the study of the prevalence of different strains of coronaviruses in different animal species in the world provides important information for the implementation of surveillance strategies, as well as epidemiological and preventive public health policies [11, 12].

## Materials and Methods

A systematic assessment of published studies reporting coronaviruses prevalence among different animal species and human, was performed. For this purpose, we used the Veterinary Science search tool at PubMed to retrieves published studies, combining different subject search terms, with temporal delimitation in years, between 2015 and 2020.

### Search strategy

The search strategy used was: veterinary[sb] AND ((“coronavirus”[MeSH Terms] OR “coronavirus”[All Fields]) AND (“one health”[MeSH Terms] OR (“one”[All Fields] AND “health”[All Fields]) OR “one health”[All Fields])) AND (“2015/04/18”[PDat] : “2020/04/15”[PDat]). From the research carried out, 190 studies were found, with the following distribution for years: in 2015 we found 18 studies (2015/18), in 2016/30 studies; 2017/41; 2018/38; 2019/34; and 2in 020 we identified 29 scientific reports.

### Inclusion criteria

The 190 references retrieved in the search process were evaluated based on titles and abstracts, and ordered by publication date. A total of 75 articles were selected because they met the inclusion criteria, and two independent reviewers carried out the selection of the articles. In the first reading step, we started by the titles, followed by the abstracts and full texts. In the selection of the titles, we included all those that presented one or more terms with a coronavirus and one health relationship. From the 75 articles 40 were selected for prevalence analysis, 42 studies for the one health analysis, 31 were used for discussion as references, and 4 articles were excluded after reading the full text.

### Data extraction

Quantitative and qualitative data extraction from the included studies was performed into four word table and an Excel spreadsheet, containing the following information: author name, year of publication, PubMed article link, article title, animal species, materials and methods, study location, and important note. During the data extraction process, information was extracted by one author and validated by a second author. Disagreements were resolved by discussion and consultation with a third author, whenever necessary.

### Quality assessment

In the evaluation of quality, an instrument adapted from the 22 criteria proposed by the STROBE Statement was used, in compliance with the principles of epidemiological investigation. This assessment aimed to classify the relevance of the articles. The One Health/ERISA evaluation scale, consisting of 15 items to evaluate the articles with regards to the existence of relevant information for the definition of novel One Health recommendations and policies.

### Data analysis

For the statistical analysis, data were stored in a predefined spread sheet file, including the authors and year of publication, number of animals and the number of infected animals.

Data were analysed using MetaXL version 5.3 software, an add-in for meta-analysis in Microsoft Excel for Windows (https://www.epigear.com/index_files/metaxl.html). The results calculated were represented in table and graphical formats. The heterogeneity across studies was evaluated by Cochrane’s Q test and I2 statistics. The calculated value of I2 allows measuring the percentage of variability due to heterogeneity, rather than chance difference or sampling error. If the value of I2 was greater than 50% and the Q test yields P<0.10, heterogeneity was considered statistically significant. The random effects model, based on DerSimonian-Laird method, which calculates the variability within and between studies, was applied to estimate the pooled prevalence and 95% CIs. The transformed double arcsine method was used for situations where the confidence limits and variance instability could appear due to any single studies with larger or small prevalence rates. The Luis Furuya-Kanamori asymmetry index (LFK index) and the Doi plot were calculated to estimate the publication bias. The presence of symmetry indicates no publication bias. The publication bias was determinate by LFK index, which can take the following assessments depending on the value obtained: no asymmetry if the LFK index is within ±1, minor asymmetry when out of the ±1 interval, but within ±2, and major asymmetry if the LFK index is beyond the ± 2 interval. A sensitivity test was calculated to provide an indication of which study is the prime determinant of the pooled result, and which is the main source of heterogeneity. The test rejects each study, one by one, in the analysis performed, so that it is possible to indicate the combined effect sizes as well as the associated heterogeneity.

## Results

### Study selection

A total of 190 studies published between 2015 and 2020 were found in our survey, with the following distribution per year: 18 in 2015, 30 in 2016, 41 in 2017, 38 in 2018, 34 in 2019 and 29 in 2020. A total of 115 studies based on the title and abstract were excluded, the remaining 75 studies were selected for the continuation of our study, and 4 studies were subsequently excluded after full reading (2 studies on bibliometrics, 1 study with lack of data and 1 study did not fit the objectives of our study). Of the 71 studies considered eligible, 42 studies were considered for systematic review and 29 studies were included in the meta-analysis (Figure 1).

**Figure.1.**
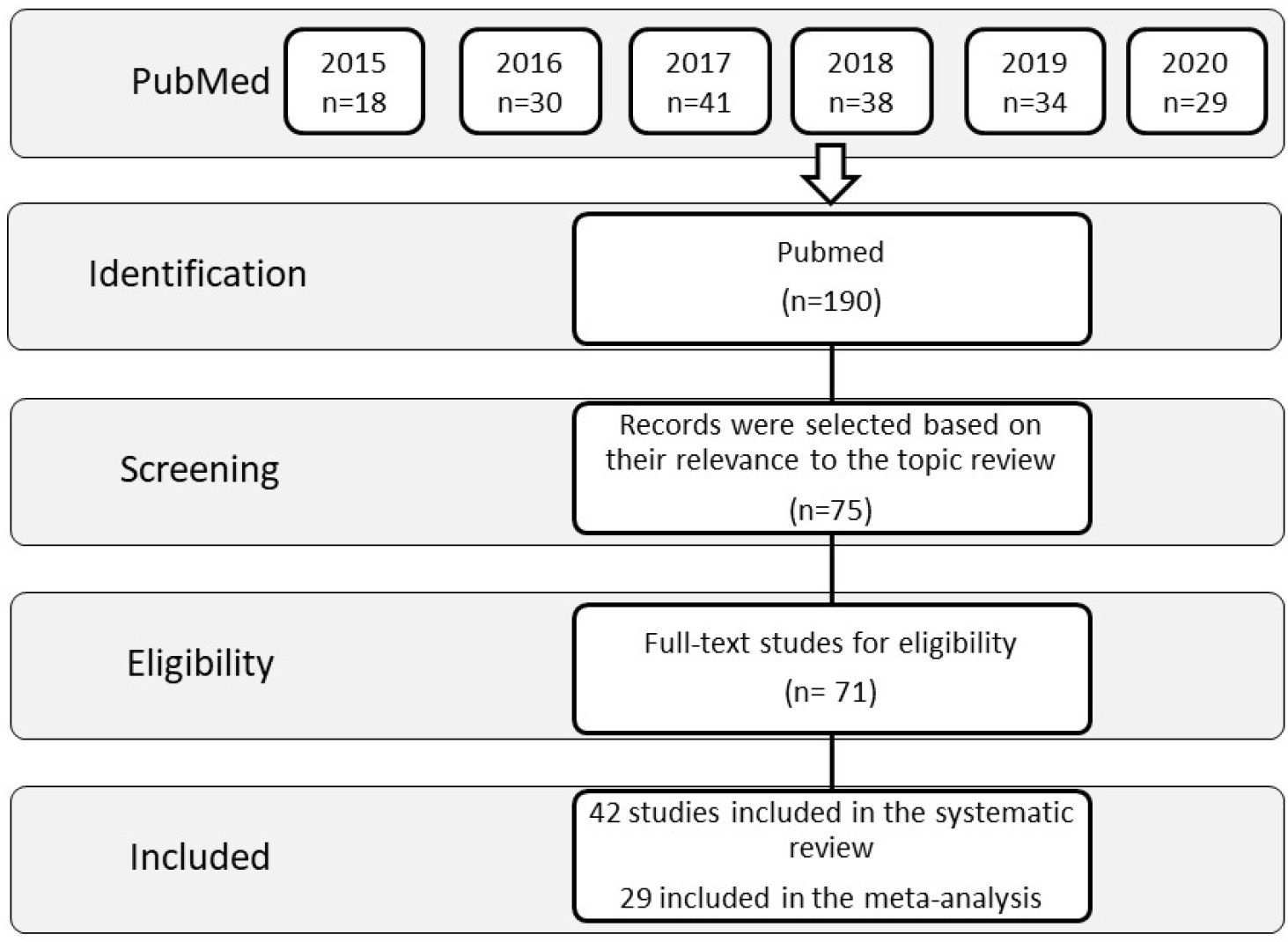
Flow chart of systematic review process.

### Quality assessment

From the one health evaluation scale, we obtained an average score of 9.33 points (62%), the scores of the articles evaluated varied between a minimum of 6 points (40%) and a maximum of 12 points (80%) in a total of 15 points (100%) possible. The following distribution of studies in relation to the average score values, obtained for each of them were: 2 studies had a score of 12 points (80%), 8 studies had a score of 11 points (73.33%), 11 studies had a score of 10 points (66.67%), 8 studies had a score of 9 points (60%), 4 studies had a score of 8 points (50.33%), 7 studies had a score of 7 points (46,67%) and one study had a score of 6 points (40%). Results are shown in Table 1. The origin of the studies carried out was as follows: three in the USA, five in China, three in Brazil, two in Saudi Arabia, one in Myanmar, one in Iran, one in Japan, one in Rwanda, one in Canada, one in Argentina, one in Qatar, one in Gulf Cooperation Council countries, one in Kenya, one in Netherlands, one in Israel, one in Italy, one in South Korea, one in Mali, one in Poland, one in Pakistan, one in Lao PDR, one in Cambodia, a joint study in Middle East (KSA, UAE, Africa: Kenya, Somalia, Sudan, Egypt), one in Sweden, one in Australia and 6 global studies (“world”) (see Table 1).

**Table 1.**
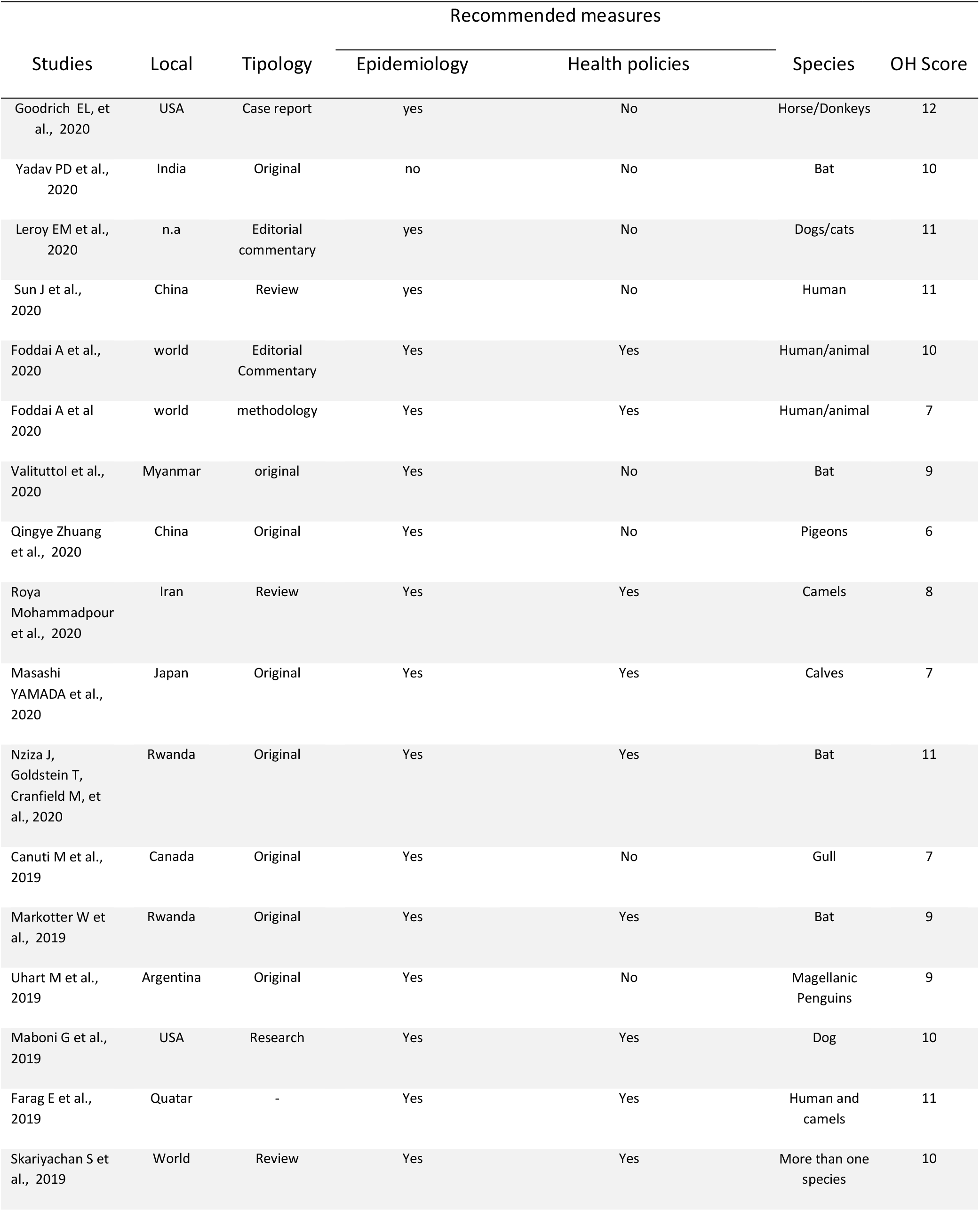
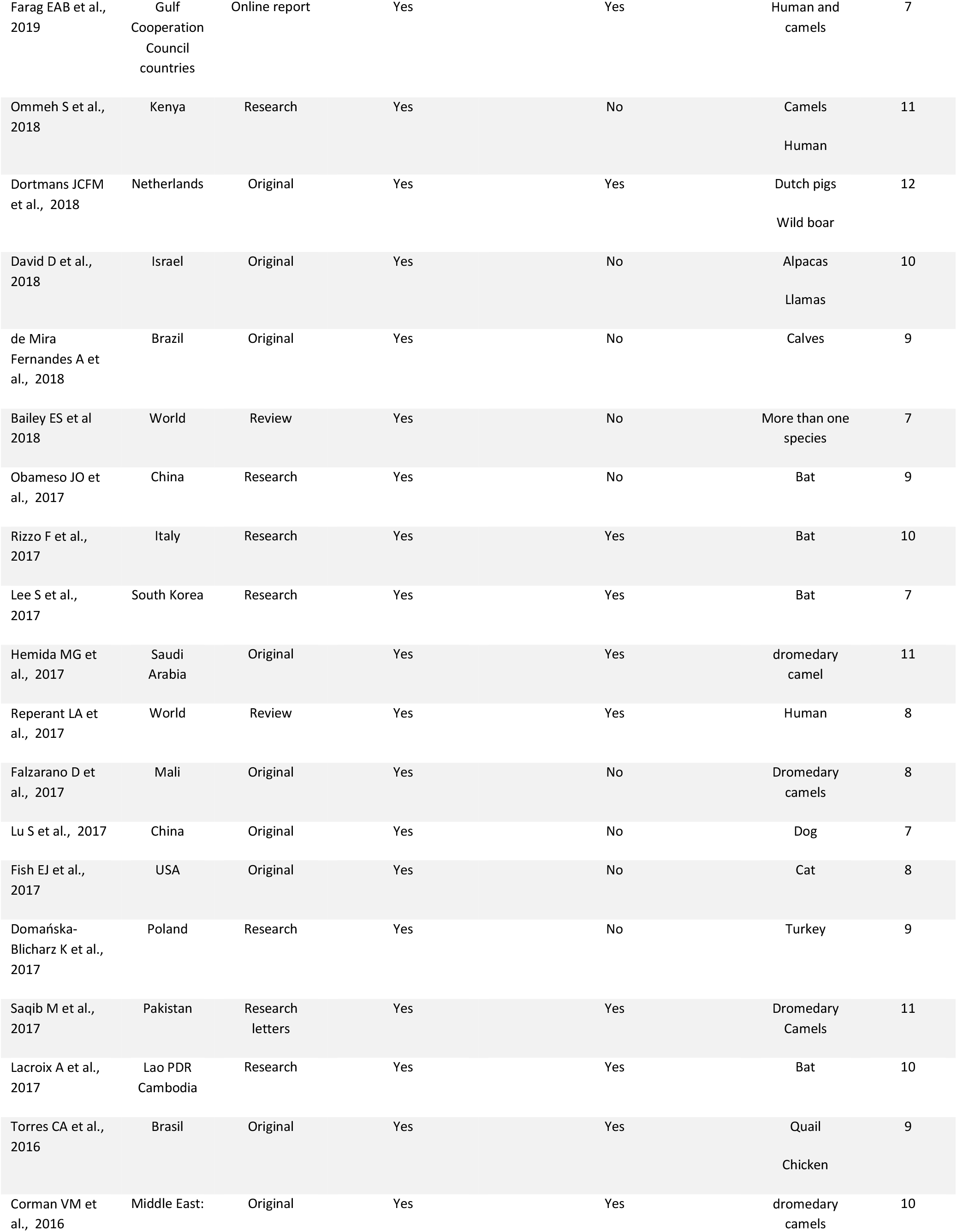
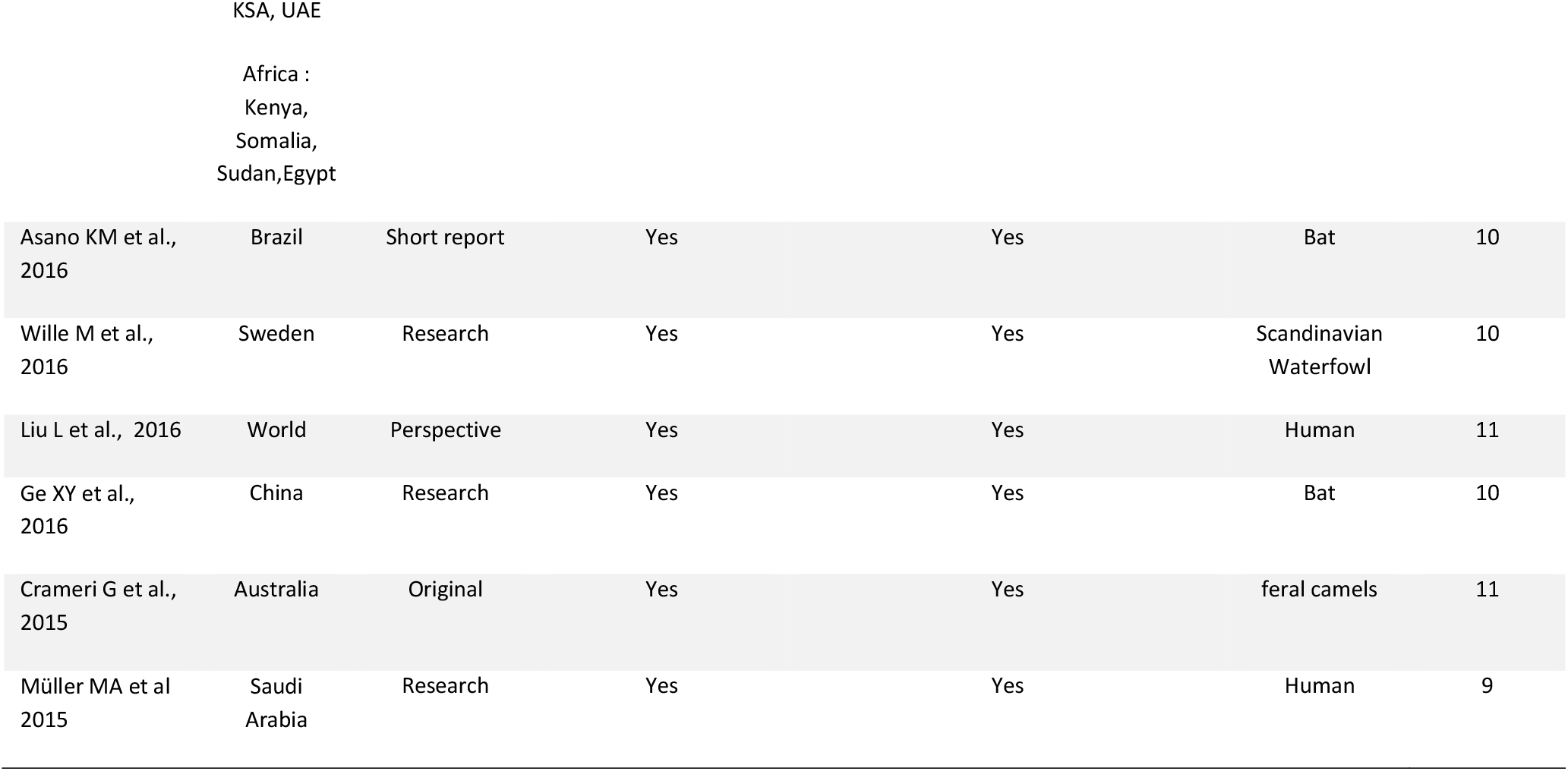
Description and quality assessment of studies with information for the definition of new One Health (OH) measures and policies:

Of the 42 studies selected for the systematic review, thirty-eight (38) contained information on epidemiological data (one study without epidemiological data), twenty-five studies (25) had relevant data for the implementation of One Health policies.

### Animal species, coronavirus strains and laboratory tests

The infected animal species described in the studies on the prevalence of different strains of coronavirus in the world were as follows: horses, donkeys, bats (various species), dogs, cats, human, bat, pigeons, camels (more than one species) calves, gull, Magellanic penguins, Dutch pigs, wild boar, alpacas, Llamas, dromedary camel, turkey, quail, chicken, Scandinavian waterfowl and feral camels. The respective identified strains of coronavirus can be found in Table 1 and 2.

### Statistics, heterogeneity and publication bias

The prevalence values of the different strains of coronavirus identified in the infected animals and described in the studies selected for the meta-analysis were projected on a world map, where the red colour indicates the maximum prevalence value and the pink signifies the lowest value, see the figure 2.

**Figure 2.**
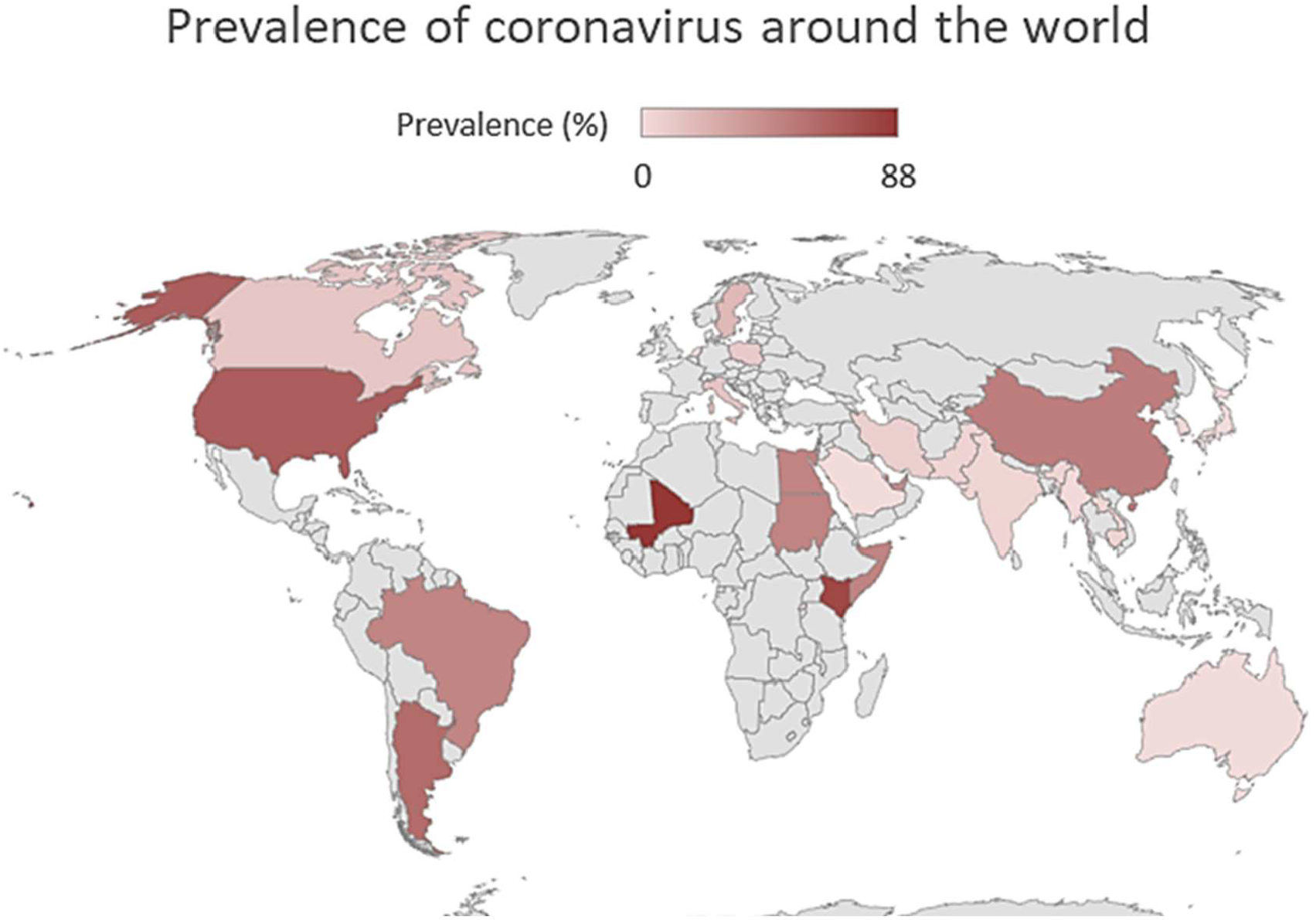
Maximum prevalence of the different coronavirus strains for each country considered in this systematic review and metanalysis.

**Figure 3.**
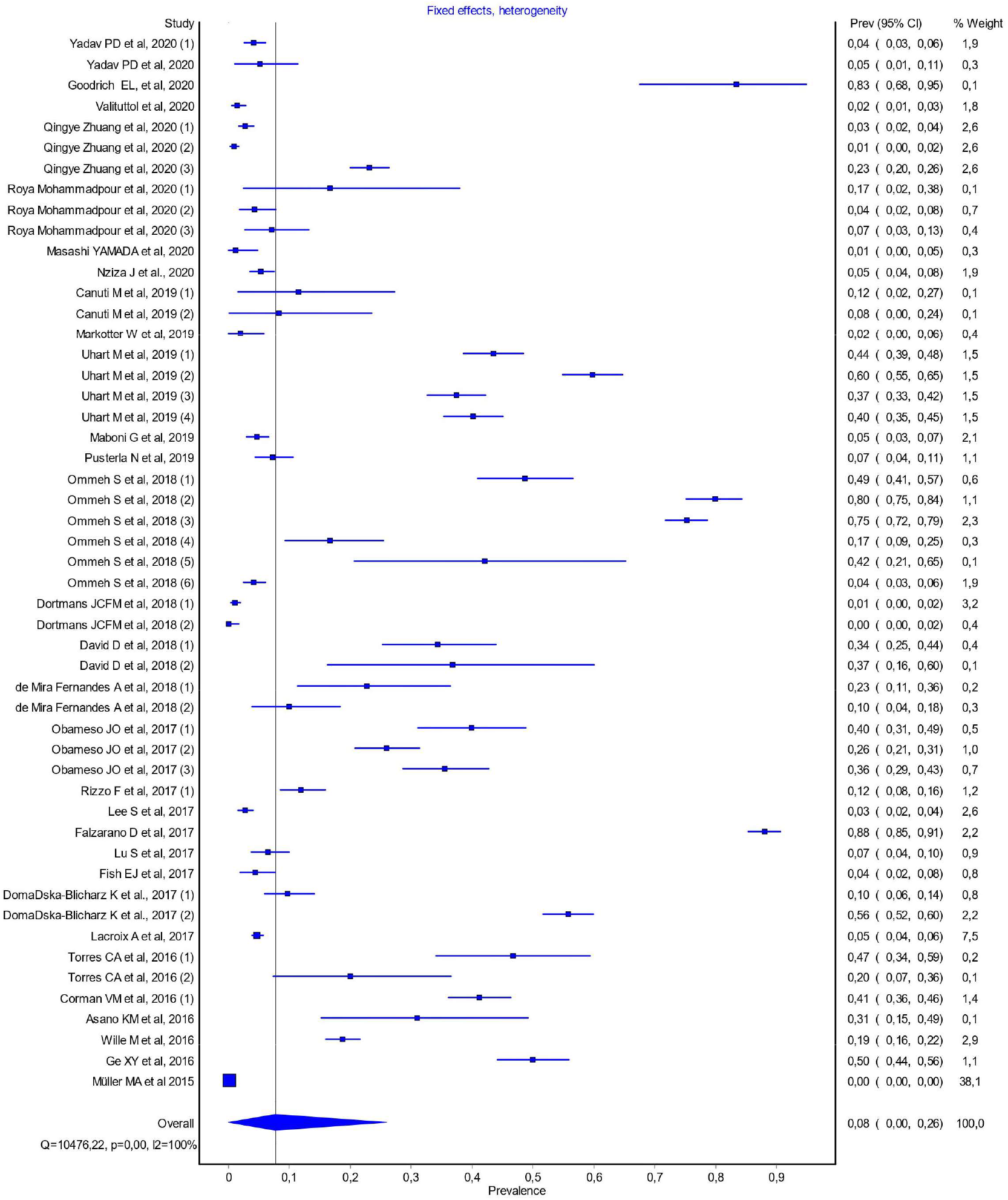
Results of metanalysis, Fixed effects, heterogeneity, Q = 10476.22, p=0.00 and I2= 100%.

The studies considered were assessed for heterogeneity through the Cochran Q test, and the results (Q = 10476.22, p = <0.001) showed that the true treatment effect is not the same across studies and variations are not caused by chance. This was confirmed by the value of the I2 test (99.2%), suggesting a statistically significant heterogeneity. The value of τ^2^(0.473), which measures the estimated variation (heterogeneity) between the effects observed in different studies also supports the fact that effect sizes vary across studies.

### Sensitivity analysis

A sensitivity analysis of the fifty-one studies was performed to evaluate the effect of each individual study on the pooled result. This was done by excluding each study step by step, one at a time (based on nine studies). The results showed that the studies of Falzarano and Müller [13,14], were the prime determinants of the pooled result.

## Discussion

### Ubiquity of coronavirus

The results obtained in this systematic review and meta-analysis demonstrate a wide variety of coronaviruses capable of infecting a very wide range of animal species, namely vertebrates. It is possible over the last few decades to see coronavirus infections in several countries on all continents, with a globally endemic virus: Sweden, Italy, Holland, Poland, USA, Canada, Argentina, Brazil, Saudi Arabia, Myanmar, Iran, Rwanda, Middle East (KSA, United Arab Emirates, Africa: Kenya, Somalia, Sudan), Egypt, Qatar, Gulf Cooperation Council countries, Kenya, Israel, Mali, Pakistan, Japan, China, South Korea Lao PDR, Cambodia and Australia (see Tables 1 and 4).

**Table 4.**
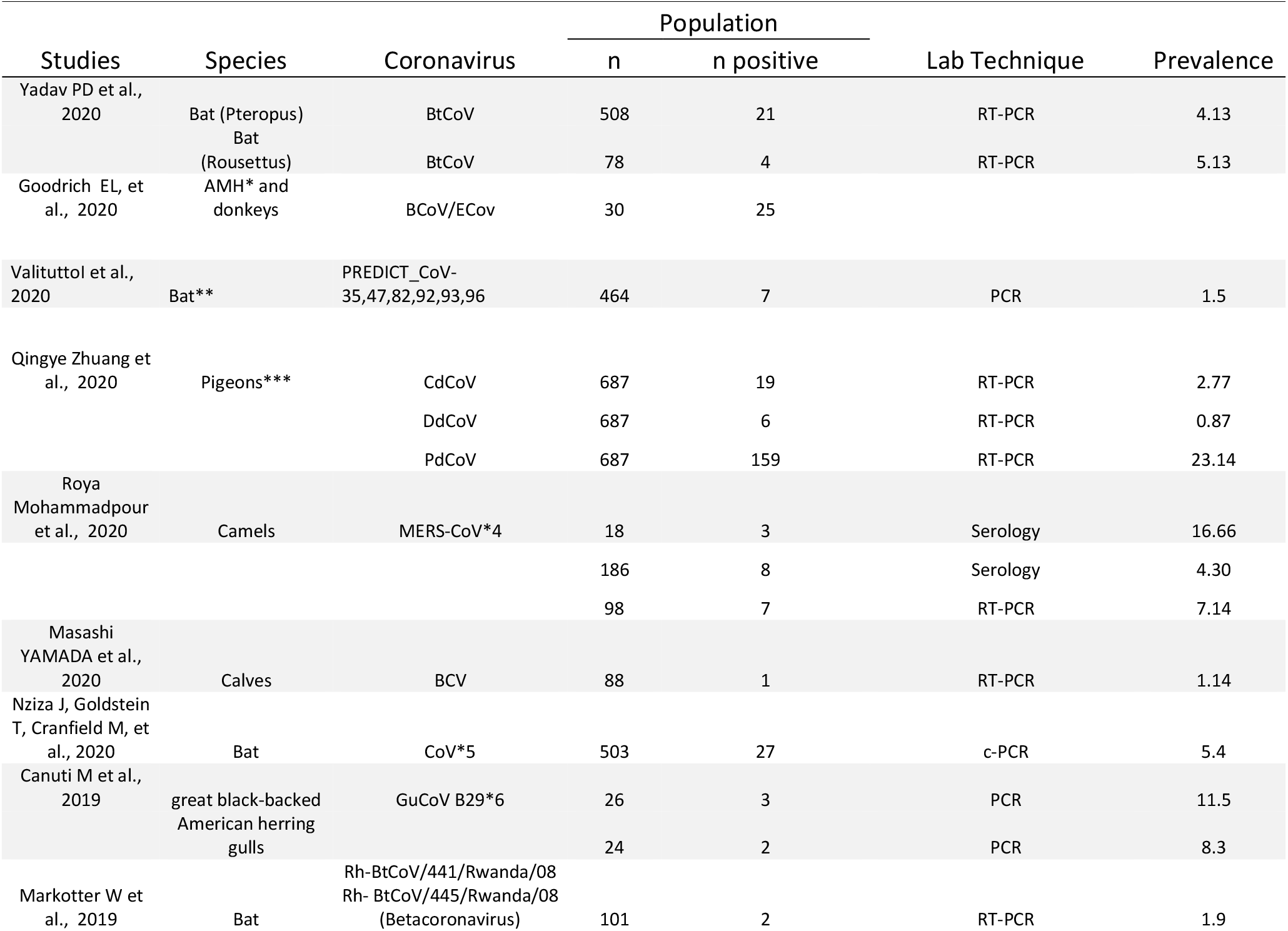
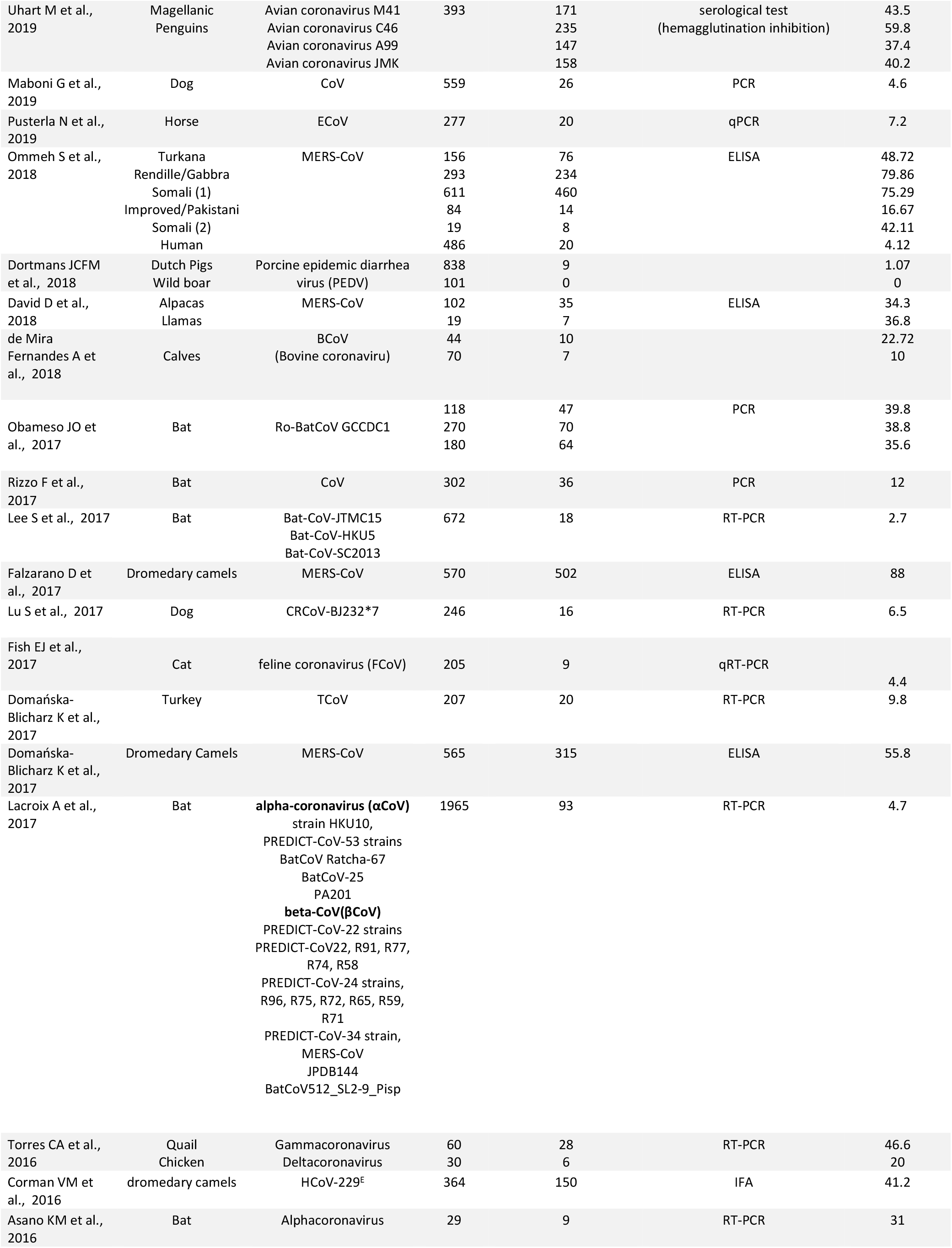
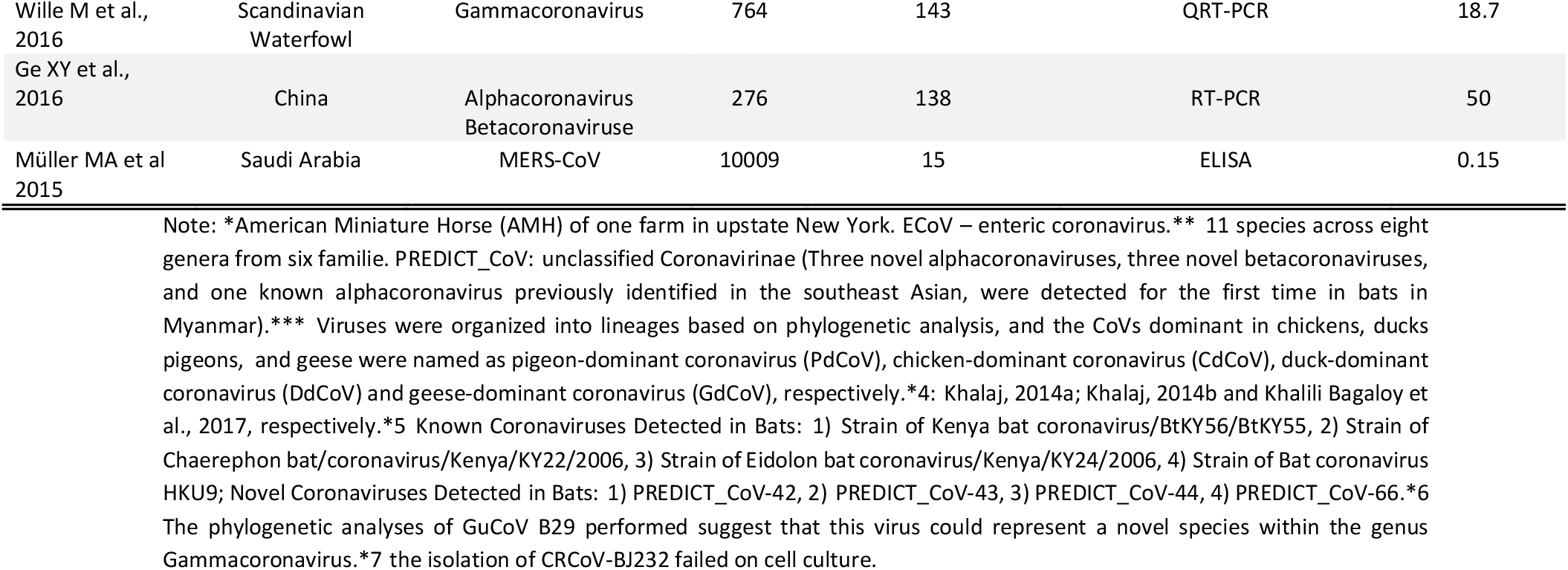
Characterization of studies according to the infected animal species and coronavirus:

### Virological diversity

If we consider the geographic regions with a prevalence greater than or equal to 0.20 (Forest plot overall; prevalence = 0.20, p = 0.00, Q = 10476.22 and I2 = 100%), we can highlight the following strains of coronavirus with the highest prevalence in this study: enteric coronavirus, ECoV (American Miniature Horse / USA) [15]; pigeon-dominant coronavirus, PdCoV (pigeons / China) [16]; Avian coronavirus M41, Avian coronavirus C46, Avian coronavirus A99, Avian coronavirus JMK (Magellanic Penguins / Argentina) [17]; MERS-CoV (camels; alpacas, llamas; dromedary camels / Kenya; Israel; Mali) [18,19]; bovine coronavirus (Calves / Brazil) [20]; Ro-BatCoV GCCDC1 (Bat / China) [21]; Gamacoronavirus, Deltacoronavirus (Quail, Chiken / Brazil) [22]; Alphacoronavirus (Bat / Brazil) [23]; Alphacoronavirus and Betacoronavirus (Bat / China) [24]. These data can be interesting in evolutionary terms, if we consider that the four human coronaviruses (HCoVs) are endemic and global respiratory pathogens. The study by Corman [25] on HCoV-229E (dromedary camels / MiddleEast: KSA and UAE; Africa: Kenya, Somalia, Sudan and Egypt) present in dromedaries infected with MERS-CoV shows that 5.6% of these animals (n = 1033) tested positive for HCoV-229E. This study was important, because MERS-CoV is an emerging strain with a zoonotic reservoir in dromedary camels and allowed us to define a hypothesis about the origin of human coronaviruses (HCoVs). The study allowed to advance that both viruses are monophyletics, with possible ecological isolation, being a descendant of camelid-associated viruses. Although HCoV-229E does not currently prove to be a risk for a global epidemic, its evolutionary history appears as a hypothesis for MERS-CoV emergence [25].

Since the appearance of the first global SARS-CoV crisis in 2003, which occurred in Guangdong province, China, with 305 cases of atypical pneumonia, the 2012 MERS-CoV and SARS-CoV-2 (COVID-19), the search for different species of animals that can be considered as a reservoir of the disease has been constant and in the case of SARS-CoV-2 inconclusive, although new evidence points to the bat [11].

### Comparative pathology studies

Three female equines of the genus Eqqus (two mares and a donkey), after returning from a show in Texas, showed signs of fever. The adult mare and the donkey showed colic and acute neurological deficits. Microscopic examination of the small intestine showed signs of severe necrotizing enteritis, with loss of epithelial lining. The lamina propria and the submucosa of the intestine of the youngest mare had multiple foci of histiocytes, lymphocytes, neutrophils and eosinophils. There was a strong diffuse, multifocal, granular to globular intracytoplasmic immunoreactivity in glandular and crypt enterocytes. In the youngest mare, lesions of severe bilateral multifocal hemorrhagic nature of the adrenal glands (Waterhouse-Friderichsen syndrome), diffuse pulmonary congestion and marked edema, generalized petechial hemorrhages in the thymus and minimal focal lymphocytic myocarditis also stood out [26].

In dromedary camels, after experimental MERSCoV infection, MERS-CoV receptors – dipeptidyl peptidase 4 (DPP4) were detected in the hair epithelial cells of the upper respiratory tract, trachea and bronchi. At the level of the pulmonary parenchyma, these receptors were detected mainly in endothelial cells and only in alveolar epithelial cells.

Contrary to that observed in dromedary camels, the MERS-CoV receptor is expressed mainly in the human lower respiratory tract (it is not expressed in a stable manner on the cell surface), with the limited expression of DPP4 in the epithelium of the human upper respiratory tract, observed in localized glands, in the submucosa of the upper respiratory tract, confirming that it is an infection of the lower respiratory tract (Widagdo W, et al., 2016) [27].

In rhesus monkeys and marmosets experimentally infected with MERS-CoV, DPP4 receptors were identified in pneumocytes type I and II, bronchial epithelial cells and alveolar macrophages. Changes in the pulmonary parenchyma led to lesions of varying degrees, including pneumonia, pulmonary edema, hemorrhage, degeneration and necrosis of pneumocytes and bronchial epithelial cells. The most prominent pathological effect observed in the lungs of rhesus monkeys was diffuse and focal eosinophilic infiltration into the thickened alveolar septum and edematous alveolar cavities, around the bronchus and between necrotic bronchial epithelial cells. In sago monkeys, diffuse and focal neutrophilic infiltration was found in edematous alveolar cavities. In both species, diffuse infiltration of numerous macrophages was observed [28].

In summary, among the general components of inflammation, fever was the predominant sign identified in mares, donkeys, rhesus and marmosets [26,28]. In the inflammatory response, there is a cellular predominance of eosinophils (chronic inflammation) and neutrophils (acute inflammation) [26,28], and lymphocytes with pycnotic and cariorretic morphology [26], with areas of necrosis both at the level of intestine, as well as at the level of the lung parenchyma [26,28]. At the level of the lung parenchyma, DDP4 expression was identified in endothelial cells, in the alveolar lining epithelium [27] and in the bronchi, as well as in alveolar macrophages [28].

### Beyond the systematic revision – Where are we with regards to the One Health Approach?

Emerging risks in agricultural and animal breeding should be addressed by specific preventive interventions. That implies a close cooperation and interaction between veterinarians, occupational health physicians and public health operators, s as to establish a worldwide strategy to expand interdisciplinary projects and better emerging risk communication policies in all aspects of health care for humans and animals, within a healthy environment. This is what the One Health Approach intends to be.

The risk of biological contamination in general is increased by a complex agricultural production process, which may persist in the “meadow to plate” chain, which favors exposure to workers in the food sector and environments. Regarding food, the first incident occurred on June 12, 2020 at the Xinfadi agricultural products market in Beijing, where SARS-CoV-2 was detected on a cutting board used to process imported salmon (Han et al., 2020). Although subsequent investigations have not been conclusive as to its origin, this particular incident raised, before authorities and consumers, some questions about frozen foods as possible carriers of SARS-CoV-2. Since the beginning of July 2020, at least nine food contamination incidents have been reported in China, where SARS-CoV-2 has been detected in imported foods, mainly packaging materials, from shrimp imported from Ecuador, and in Shenzhen, in Guangdong province on August 12, 2020, on the surface of frozen chicken originating in Brazil, which became the first known case in which the new coronavirus was detected in real samples of imported foods [29].

. It is worth considering the possibility that the food cold chain may promote contamination, because laboratory studies [30] have shown that SARS-CoV-2 remained highly stable under refrigeration, at 4° C, and in freezing conditions, from -10 to -80 °C in fish, meat, poultry, and pig skin for 14–21 days. In a controlled laboratory study [31], the persistence of SARS-CoV-2 in chilled salmon, frozen chicken and pork for 21 days was examined. The study showed that SARS-CoV-2 titers remained virtually constant, and the inoculated viruses maintained their infectivity both in the refrigerated product (4°C) and in the frozen samples (−20 °C and -80 °C). In a previous study, researchers presented evidence that SARS-CoV-2 can remain quite stable in pig skin for 14 days, at 4 °C [32].

The most recent investigation, in an experimental context, points to the new coronavirus remaining up to 72 hours in plastic and stainless steel with temperatures of around 20° and humidity of 40% [33, 34]. Other multiple investigations have already reported that SARS-CoV-2 and other coronaviruses are able to remain on surfaces such as metal, glass, PVC, Teflon, and other materials, for several days [35,36,37,38].

At the interface between the health of humans, animals and the ecosystem, host receptor recognition is a determinant for virus infection. Recently (2020) Li [39] conducted sequence and structural analyses of angiotensin-converting enzyme 2 (ACE2) from different species, which sheds some light on cross-species receptor usage of SARS-CoV-2. Citing the authors, all these analyses raise an alert on a potential interspecies transmission of the virus and propose further surveillance in the diverse animal populations.

## Conclusions

### A true One Health approach is urgent

This systematic review and meta-analysis of the prevalence of the coronaviruses worldwide highlights the great biological diversity of these agents, as well as their ability to infect a wide variety of species. The latest most important epidemiological crises, which occurred in 2003 (SARS-CoV-1), 2012 (MERS-CoV) and 2019 (SARS-CoV-2, COVID-19) alert to the potential epidemic risk of this infection, which causes severe atypical pneumonia and several other systemic dysfunctions.

The wide variety of infected animal species or natural sources of the coronaviruses require a crosscutting and multidisciplinary approach. It is essential to put in place a One Health approach to this public health problem and to take advantage of the experience from the collaboration between Human and Veterinary Medicine. The concept of One Health has become increasingly important. However, there is much to do. It is essential to enhance the structuring of new One Health policies that allow the creation of epidemiological surveillance programs and the creation of advanced training courses in this new area of intervention that brings together human, animal and environment health, in order to prepare human resources to fight the next pandemic. The SARS-CoV-2 (COVID-19) pandemic only showed our lack of preparedness and responsiveness at the global level. Will we be better prepared for the next one?

## Data Availability

the author declares that all data are available in the current manuscript.

## Authorship contribution

Ricardo Faustino: conceptualization, data management, metanalysis, investigation, methodology, validation, writing - original draft, review, editing, supervision. Paulo Sargento: investigation, methodology, validation, review and editing. Maria do Céu Costa: conceptualization, investigation, methodology, validation, Writing and Review. Mónica Teixeira: conceptualization, investigation, methodology, validation, writing and review. Miguel Faria: Investigation, methodology, validation, review metanalysis, writing and editing. Filipe Palavra: conceptualization, investigation, methodology, validation and review.

## Acknowledgements

The present study was supported by the Research Group in Health Sciences and Technologies - NICiTeS, Ribeiro Sanches Higher School of Health, Polytechnic Institute of Lusophony (IPLuso) Lisbon, Portugal

